# Clinical Utility of EsoGuard® as an Efficient Triage Test for Diagnosing Barrett’s Esophagus in On-Duty Firefighters

**DOI:** 10.1101/2023.08.16.23294176

**Authors:** Rachelle Hamblin, Victoria T. Lee, Brian J. deGuzman, Suman Verma, Lishan Aklog

**Affiliations:** Department of Medicine, CHRISTUS Health, San Antonio, TX, USA; PAVmed Inc.; New York, NY, USA; Lucid Diagnostics Inc.; Lake Forest, CA, USA

## Abstract

**Background:** Firefighters have frequent exposure to compounds shown to increase risk of esophageal neoplasia. EsoGuard® (EG) is a DNA biomarker assay that can be utilized with efficiency and high tolerability as a triage to endoscopy for diagnosis of patients with Barrett’s Esophagus (BE), a known precursor to esophageal adenocarcinoma (EAC). This diagnostic tool may facilitate disease testing among busy at-risk firefighters.

**Methods:** Retrospective analysis of prospectively collected clinical utility (CU) data for use of EG as a triage to more invasive endoscopic evaluation. EG was performed on esophageal cell samples collected with the nonendoscopic EsoCheck® (EC) device during two large cancer and pre-cancer screening events for firefighters in San Antonio, TX, in January 2023. CU was evaluated by provider impact assessment.

**Results:** 388 firefighters were identified for EG testing, of which >99% (385/388) successfully completed EC cell collection. Over 96% (372/385) of tests had binary results; the remaining <4% failed analysis due to insufficient DNA. The EG positivity rate was 7.3% (28/385), all of whom were referred for specialist and upper endoscopy evaluation. Among those who tested negative, none were referred for further diagnostic workup. This represented a 100% concordance between EG results and physician management decisions.

**Conclusions:** This study capturing real-world data on use of EG in a population of firefighters demonstrates its ability to test many individuals rapidly and efficiently in a well-tolerated fashion, and reliable use of the test to triage individuals prior to pursuing more invasive and time-consuming diagnostic approaches.

## 1. Introduction

Esophageal adenocarcinoma (EAC) is the most common cancer of the esophagus in the United States (US), with an incidence that has been increasing over the last 40 years, particularly in white males, for whom the incidence has gone up more than 6-fold since the 1970’s.[1-3] National statistics estimate there were 20,640 new cases of esophageal cancer, mostly adenocarcinomas, in 2022, with estimated 16,410 deaths.[4] Despite advances in chemotherapy, radiotherapy, and surgical therapy, the prognosis for EAC remains poor, with a 5-year survival rate of only 20%.[4-5] Barrett’s Esophagus (BE) is a direct precursor to EAC and has well defined risk factors which have been utilized in published guidelines to establish recommendations for screening, including those from the American College of Gastroenterology (ACG) and other Gastroenterological societies.[6-7] This is because, contrary to the lethality of EAC, BE when detected early, can be treated using highly effective endoscopic approaches such as radiofrequency or cryotherapy ablation with upwards of 80% success rates.[8-10] However, most individuals, including many of those deemed at high risk for disease based on risk factors, do not undergo recommended screening endoscopies;[11] indeed, less than 20% of patients in the U.S who are diagnosed with EAC have any preceding diagnosis of BE. [12] Barriers of conventional screening endoscopy may include the need for specialist referral and sedation, and patient concerns about invasiveness, procedural complications, and logistics of scheduling.[13] Recently, to bridge this gap, non-endoscopic cell collection devices such as EsoCheck® (EC) have been developed, and when paired with a biomarker test such as EsoGuard® (EG), have been endorsed by both the ACG and American Gastroenterological Association (AGA) as a reasonable alternative to upper endoscopy (UE) for BE screening.[6-7]

One population of particular interest for BE/EAC screening are firefighters, who by nature of their occupation have ongoing exposure to multiple suspected and known carcinogenic agents, such as (but not limited to) formaldehyde, benzene, asbestos, and polycyclic aromatic hydrocarbons. In July of 2022, firefighting was designated a Group 1 carcinogen by the International Agency for Research on Cancer (IARC). [14] Additionally, a recent pilot study from France suggests a link between specific occupational exposures (including those common to firefighters, such as asbestos, hydrocarbons, etc.) and esophageal cancer.[15] Excess incidence of cancers of the digestive tract are observed within the firefighter population, namely esophageal and colorectal malignancies.[16-17] As such, firefighters have the potential to significantly benefit from an easily accessible, minimally invasive, non-endoscopic screening test for BE. EC/EG can be implemented in nearly any outpatient setting and used to triage patients prior to pursuing more invasive time-consuming workup; this approach broadens access to testing among elevated-risk individuals while also focusing endoscopy resources on those with highest probability of disease. The goal of improved BE detection is to establish surveillance or schedule treatment, and ultimately reduce EAC mortalities by halting disease progression in a pre-neoplastic stage. [6]

This study evaluates clinical utility data captured from San Antonio firefighters who underwent clinically directed EsoGuard testing as part of two large cancer and pre-cancer screening events in January 2023. Clinical utility was assessed using provider impact assessment. To our knowledge, this was the first incidence of any large-scale screening for BE/EAC in U.S firefighters to date.

## 2. Materials Methods

### 2.1 Population

Two large health fairs for cancer and pre-cancer screening among San Antonio firefighters were organized by over 40 community volunteers, including community physicians and other health care professionals in January of 2023. The events occurred over the course of two weekends (January 14-15, and January 28-29). Resources and support for skin cancer screening were provided by Mollie’s Fund (Mollie Biggane Melanoma Foundation), and Lucid Diagnostics Inc. provided staffing support for screening of BE. Firefighters underwent clinically directed EG testing on esophageal cell samples collected using EC (EC/EG). The decision to test any given individual was based on the physician’s assessment of risk, as determined by presence of ≥3 risk factors for BE/EAC, including known occupational risk, history of gastroesophageal reflux disease (GERD) and/or chronic heartburn symptoms, tobacco smoking, obesity, male sex, white race, age >50 years, and family history of BE/EAC.

Patients identified to have “red flag” symptoms such as dysphagia or escalation of pre-existing symptoms were automatically referred for specialist evaluation (Gastroenterology) and diagnostic UE without EC/EG and are not included in the scope of this study. All other patients were educated by the physician about BE/EAC, risk factors, and the EC/EG technology. They were given the option to undergo EC/EG testing, and those who agreed were then passed to trained nurse practitioners for cell collection. This was performed according to the EC device Instructions For Use (IFU), and samples sent to the central lab for analysis (LucidDx Labs, Inc, Lake Forest, California)

### 2.2 Ethical Considerations

The study was conducted according to the guidelines of the Declaration of Helsinki and approved by the WCG Institutional Review Board (study number 1350589, approved on 03-March-23). Given the retrospective nature of the analysis, and satisfactory plan for protecting patient identifiers from improper use and disclosure, patient informed consent was waived.

### 2.3 EsoCheck® and EsoGuard®

EsoCheck® (EC) is an FDA cleared, non-endoscopic cell collection device (**Figure 1**) designed to circumferentially sample cells from a targeted region of the esophagus (**Figure 2**); EsoGuard® (EG) is a laboratory developed test (LDT) performed in a Clinical Laboratory Improvement Amendment (CLIA) certified and College of American Pathologists (CAP) accredited lab that utilizes set of genetic assays and algorithms which examines the presence of cytosine methylation at 31 different genomic locations on the vimentin (VIM) and Cyclin-A1 (CCNA1) genes. EG results are reported in a binary fashion (positive or negative) indicating presence or absence of sufficient methylation changes to suggest diagnosis of BE or disease along the BE to EAC progression spectrum. EG has been clinically validated in a developmental study published in 2018 and shown to have a >90% sensitivity and >90% specificity in non-endoscopic detection of BE or EAC.[18]

**Figure 1.**
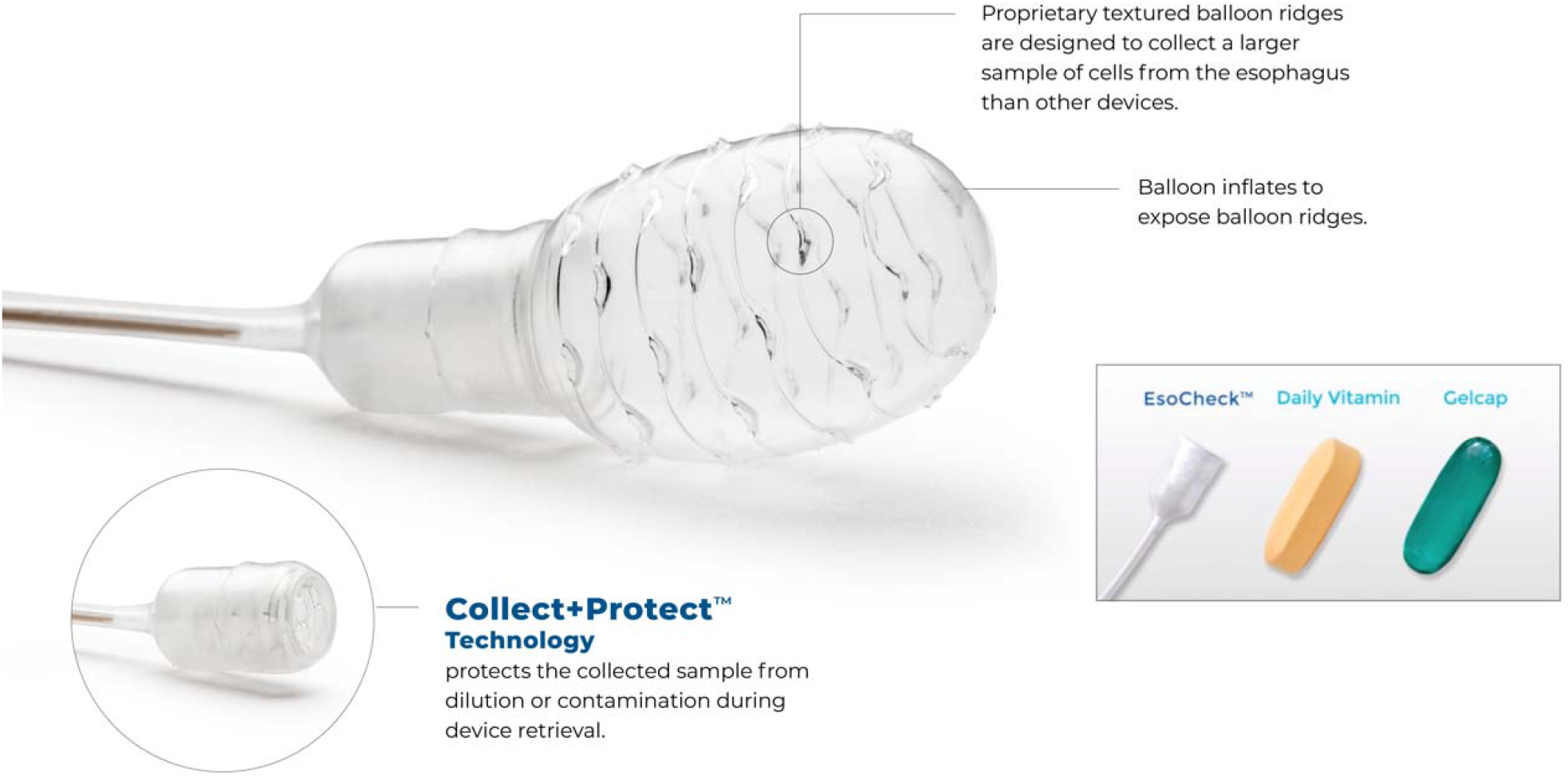
EC administration is a simple, non-invasive, non-endoscopic, office-based procedure that can be performed by a variety of healthcare providers including physicians, nurse practitioners, physician assistants, nurses, or other trained personnel usually in less than 5 minutes and without sedation or significant pre-procedure preparation.

**Figure 2.**
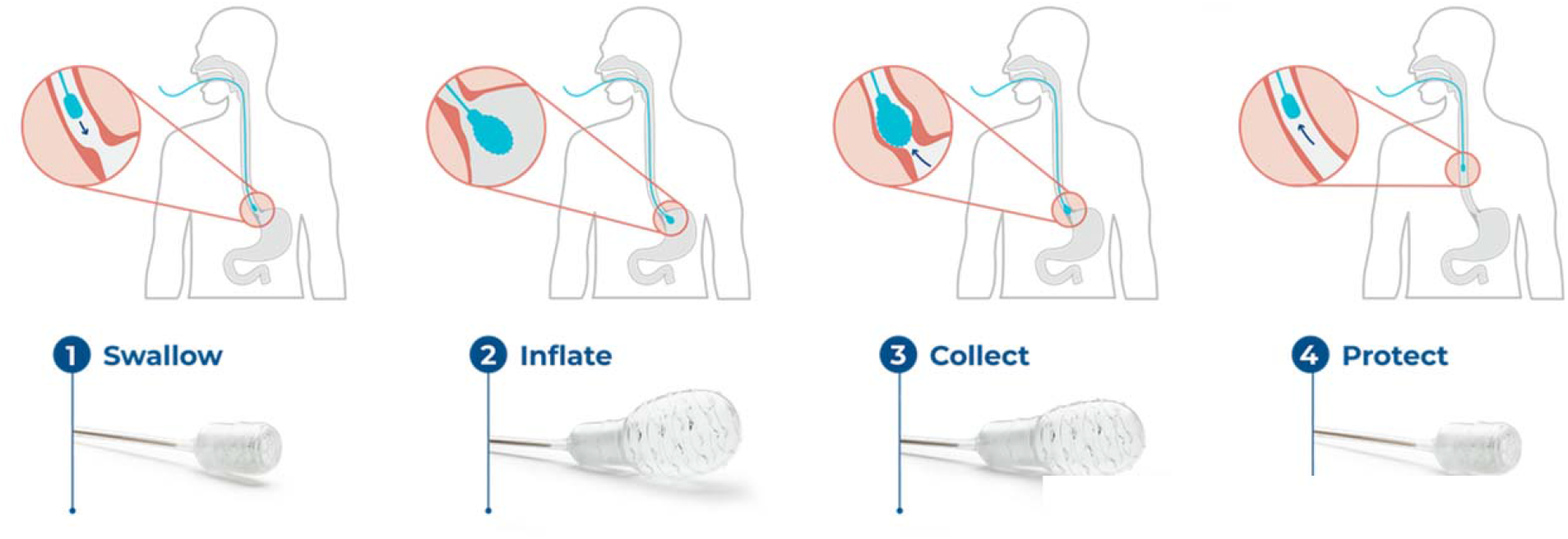
Non endoscopic cell-collection devices paired with a biomarker test (e.g., EC/EG) are deemed an acceptable alternative to UE to screen for BE, according to the 2022 ACG guidelines and AGA clinical practice updates for screening in BE.[6-7]

### 2.5 Follow-up

EG results were available within two weeks of cell collection. All EG results were reviewed by the ordering physician, and the decision then made on whether referral for specialist evaluation and UE would be provided. All negative results were conveyed by a follow-up letter, with an explanation of the results and recommendations for ongoing care (including a proposed monitoring plan for individuals deemed to be particularly high risk, based on the number of risk factors). All positive results were communicated by the ordering physician directly to the patient, with an explanation of the results and further recommendations for care. Further care coordination included referral to either an experienced gastroenterologist, general surgeon, or bariatric surgeon who could then perform the necessary confirmatory UE (with biopsies as clinically indicated), counseling, and develop a treatment or surveillance plan for positive results. Hard copies of positive EG results were also given securely to patients as well as providers to ensure continuity of care.

### 2.6 Data Collection, Provider Impact Assessment, and Statistical Analysis

This is a retrospective analysis of prospectively collected data on the clinical utility of EC/EG in the San Antonio firefighter population; data was collected during two large health fairs for cancer and pre-cancer screening in January of 2023. A retrospective chart review was performed, and a limited data set collected, consisting of patient demographic information, EG results, and patient management decisions – namely whether the patient was referred by the ordering physician for specialist evaluation and UE. Provider impact assessment and therefore the EG clinical utility evaluation was based on the patient management decisions. Most BE/EAC risk factors obtained during the health fair were by patient self-report (via questionnaire) and not validated against medical records at the time of testing; thus, they were not included within the scope of this analysis.

All data was compiled in an Excel file. As this is not a hypothesis-driven study, no statistical software was utilized for data analysis, and calculations were performed with Excel. The results for continuous variables are shown as medians with interquartile range (IQR). Data from patient test results and outcomes are presented as numbers and percentages. No comparative tests were performed.

## 3. Results

### 3.1 Patient Characteristics

A total of 388 San Antonio firefighters were ordered for EG testing over two weekends. Of these, 385 (99.2%) successfully swallowed the EC device to provide cell samples. The remaining three were unable to tolerate the cell collection and could not provide DNA for the EG assay; they were excluded from study analysis.

An overview of testing numbers and basic patient characteristics is provided in **Table 1**. Males accounted for 93.0% (358/385) of the tested population, and females accounted for the remaining 7.1% (27/385). The median age was 41.5 years [IQR 14.45]. Given that retirees did not participate in the health fair (participants were active-duty firefighters only), most patients (76.6%; 295/385) were <50 years old, but less than 10% (9.6%; 37/385) younger than 30 years. The median age for the males and females were similar (41.5 years old [IQR 14.45] and 42.3 years old [IQR 12.65], respectively).

**Table 1.** Testing Numbers and Patient Characteristics.

|  | n |  | % |
| --- | --- | --- | --- |
| Firefighters participating in EsoCheck/EsoGuard BE/EAC screening | 388 |  | 100 |
| Firefighters unable to swallow EsoCheck (i.e., unable to provide cell samples for EsoGuard) | 3 |  | 0.1 |
| Firefighters who successfully swallowed EsoCheck | 385 |  | 99.2 |
| **Full Analysis Cohort** | **385** |  |  |
| Median age (years) | **41.5** | **IQR: 14.5** | **Min, max: 21.6, 77.5** |
| Male | 358 |  | 93.0 |
| Median age (years) | 41.5 | IQR: 14.5 | Min, max:<br>21.6, 77.5 |
| Female | 27 |  | 7.1 |
| Median age (years) | 42.3 | IQR: 12.7 | Min, max:<br>29.6, 63.9 |

### 3.2 EsoGuard Results

There were 372 patients who received binary EG results (96.6% successful analysis rate) and only 13 cell samples had insufficient DNA quantity for EG analysis (‘quantity not sufficient,’ QNS; 3.37%). Twenty-eight patients tested positive (7.3%), while 344 patients tested negative (89.4%). See **Table 2** for details.

**Table 2.** EsoGuard® Results.

| Result | n | % |
| --- | --- | --- |
| **Total** | **385** |  |
| **Positive** | **28** | **7.3** |
| Male | 26 | 92.9 |
| Female | 2 | 7.1 |
| **Negative** | **344** | **89.4** |
| Male | 319 | 92.7 |
| Female | 25 | 7.3 |
| **DNA Quantity Not Sufficient (QNS)** | **13** | **3.4** |
| Male | 13 | 100.0 |
| Female | 0 | 0 |

Among the EG positive patients, two (7.1%) were female and the remainder were male (92.9%. 26/28;). All QNS test results were from male patients. The EG positive results were analyzed based on characteristics of patient sex and age (**Table 3**), although the sample size of females was too small to make inferences. EG positivity had a trend towards increasing in older age-groups, notably in patients age ≥50. Although 50% of the patients aged ≥70 years ended up testing EG positive, the sample size was again too small to make inferences.

**Table 3.** EsoGuard® Positive Results by Age and Sex.

| Characteristic | n<br>EsoGuard<br>(+) | Total n | Positive<br>rate (%) |
| --- | --- | --- | --- |
| Male sex | 26 | 358 | **7.3** |
| Female sex | 2 | 27 | **7.4** |
| Age <30 years | 0 | 37 | **0.0** |
| Age <50 years | 15 | 295 | **5.1** |
| Age 50 years or greater | 13 | 90 | **14.4** |
| Age 70 years of greater | 3 | 6 | **50.0** |

The average age of the EG positive patients was 44.6 years old, with the youngest being 30.9 and the oldest being 77.5 years old. Among the EG positive patients, 85.71% (24/28) self-reported a history of either GERD or regular heartburn symptoms.

All 28 EG positive patients were referred by the ordering provider for UE evaluation. None of the 344 EG negative patients were referred for further diagnostic work-up. The 13 patients with QNS EG results were offered the opportunity to re-test, all of which elected to do so. Taken together, there was a 100% concordance between the EG assay result and the ordering provider’s management decision for the patient.

## 4. Discussion

In the U.S and other Western countries, the most common type of esophageal cancer is EAC, which is known to arise from the pre-malignant condition of BE.[19] Chronic GERD has long been associated with the development of BE and EAC, with additional demographic and lifestyle characteristics such as male sex, older age (>50 years), white race, tobacco smoking history, obesity, and family history being well-described and quantified as risk factors.[20] As such, high-risk patients with multiple risk factors are recommended for BE screening, followed by recommendations for long-term surveillance or treatment of those diagnosed with disease. While not a risk factor traditionally linked with BE or EAC, a growing body of literature suggests that firefighters are at increased risk. One of the earliest suggestions of an association between firefighters and esophageal malignancy arose from a registry-based case-control study published in 2007. Based on records of 3,659 California firefighters, there was evidence that firefighting could be a risk factor for development of esophageal cancer, with an odds ratio of 1.48 (95% CI 1.14– 1.91).[21] In a pooled cohort of U.S firefighters from San Francisco, Chicago, and Philadelphia, evaluating mortality and cancer incidence from 1950 to 2009 (later with updated mortality data through 2016), the standardized mortality ratio (ratio of observed to expected number of deaths) was 1.31-1.39 for esophageal cancer; the standardized incidence ratio (ratio of observed malignancies to the expected number of cases estimated using U.S incidence rates) was 1.62.[16-17] Similar results were found by the National Institute for Occupational Safety and Health (NIOSH) in their study of California firefighters from 1988-2007 with an odds ratio of 1.6 for esophageal cancer.[22]

This study is the first to evaluate use of a nonendoscopic cell collection device (EC) paired with a biomarker test (EG) to evaluate a high-risk firefighter’s population for BE. Unfortunately, most patients who meet guideline recommendations for BE screening are not undergoing testing.[23] Over 20% of surveyed GERD patients reported fear of discomfort from screening endoscopy as a barrier, along with logistical and accessibility concerns such as identifying a procedure location, scheduling the procedure, wait time/out of work time, etc. [13] EG, when used to analyze samples collected using EC, was developed to address this diagnostic gap, by providing an easy, in-office alternative to screening endoscopy. Although not intended a replacement for UE to investigate patients with alarm symptoms or to diagnose other esophageal pathologies, EC/EG could be a viable triage tool for identifying patients at high vs. low likelihood of disease, and this utility has already been recognized by the ACG and AGA.[6,7] The EG assay has a published sensitivity of >90% for detecting disease along the full BE to EAC disease spectrum, which is comparable or even superior to that of cervical pap smears (50-80% sensitivity), which is has long been a widely accepted approach to cervical pre-cancer detection.[18, 24]

In the San Antonio firefighter population, EC was successfully swallowed in >99% of patients, and there were no complications reported. Binary EG test results were available in over 95% of patients, which remains well within standards set within the Central Lab’s (Lucid Dx) CLIA requirements. The tolerability of EC/EG is denoted by the willingness of all patients with QNS results to schedule a repeat cell collection, although the repeat testing had not been performed by the time of data analysis. Indeed, a major benefit of EC/EG for the firefighters was the ability to undergo testing while on-shift. The average cell collection time is shorter than five minutes, and as discussed previously can be performed in most out-patient settings, including at the fire training academy where the health fairs/cancer and pre-cancer screening events were conducted during both January weekends. All tested individuals were able to return to duty immediately after completion of EC. This is in stark contrast to screening upper endoscopy, which requires an endoscopy suite, sedation, and post-sedation recovery time. As seen in this San Antonio firefighter testing experience, EC can be administered across large numbers of patients in an efficient and well-tolerated fashion with circumferential cell sampling and high DNA yield. **Table 4** provides a summary of the EC characteristics (compared with upper endoscopy) and patient experience variables which the authors believe contributed to the successful testing of nearly 400 firefighters over only four days.

**Table 4.** Patient Experience Variables: EsoCheck® vs. Screening Upper Endoscopy.

|  |  | **EsoCheck®: nonendoscopic esophageal cell collection** | **Screening Upper Endoscopy** |
| --- | --- | --- | --- |
| **Patient experience variable** | Pre-procedure NPO requirement | **2hrs** | **6-8hrs** |
|  | Requires a specialist physician to perform? | **No** | **Yes** |
|  | Requires a specialized test location (e.g., endoscopy suite or procedure room)? | **No** | **Yes** |
|  | Anesthesia or sedation required? | **No** | **Yes** |
|  | Education/consent time | **15min** | **15min** |
|  | Test time | **Average 3min** | **Approximately 30min<sup>26</sup>**<br>(dependent on findings and number of biopsies performed) |
|  | Post-procedure time on-site | **5min** | **Up to 2hrs<sup>26</sup>**<br>(post-sedation monitoring) |
|  | Total patient time commitment | **<30min** | **>2hrs** |
|  | Firefighters able to return to shift after the procedure? | **Yes** | **No** |

The EG positivity rate within the tested population (7.3%) is consistent with expected BE prevalence rates from the literature (5-15%).[25] The presence of self-reported GERD and/or chronic heartburn symptoms in >80% of tested individuals, and higher EG positivity rate among those aged 50 and older is consistent with what would be expected based on established risk factors for disease. The provider impact assessment demonstrated a 100% agreement between positive EG results and the physician decision to refer the patient for specialist evaluation and UE. There was also 100% agreement between negative EG results and the physician decision not to refer the patient for UE. This demonstrates reliable use of EC/EG by the ordering physician as a triage tool to inform decision-making on the next steps in patient management. Only EG positive patients were sent for confirmatory evaluation, whereas physician confidence in the high sensitivity of EG spared the negative patients from a more-invasive diagnostic test. Firefighters in particular stand to strongly benefit from BE testing that is efficient and allows for minimal to no “out of service time” (enabling them to respond to calls if needed), while providing their physicians with information to guide care. These initial steps of patient education, (non-endoscopic) screening, and establishing care with a gastroenterologist or other relevant specialist is critical, as once diagnosis has been confirmed with UE, existing guidelines provide clear recommendations for BE surveillance (non-dysplastic disease) and treatment (dysplastic BE), to minimize risk of progression to cancer.[6,7]

This study provides the first real-world example of how EC/EG can facilitate diagnosis of a cancer precursor condition in the elevated-risk firefighter population. The benefit of a triage tool such as EC/EG is to improve overall access to testing, while focusing utilization of the expensive and more invasive UE procedure on the highest risk patients. We recognize certain limitations within our study, including the observational nature and single-provider experience. Additionally, not all tested patients met ACG guideline criteria or even AGA recommendations for BE screening. The ordering physician deemed firefighting/occupational exposure to be one of 3 (or more) necessary risk factors to warrant testing, although it is not currently recognized as such within any society guidelines. Finally, given that most risk factors including obesity, family history of BE/EAC, and tobacco smoking history were patient-reported, these were excluded from current analysis, as they were unvalidated against patient medical records, and anecdotally patients were frequently observed to under-report their obesity status.

This study was also not intended to validate the EsoGuard assay performance, but rather to evaluate the clinical utility of the test within a population of high-risk group of individuals. Future directions for study would include evaluation of an even larger and geographically diverse population of firefighters, more comprehensive collection of individual risk factors, and longitudinal follow-up which could be powered for subgroup analysis. While the current study only tested active-duty firefighters, future studies may benefit from testing of retirees who have the longest duration of occupational exposure and would be at highest risk. However, despite the above limitations, we present the largest experience to date of BE/EAC screening in firefighters utilizing nonendoscopic strategies. Findings were reassuring that ordering providers can and will consistently utilize EC/EG as intended - namely as a tool for triaging patients prior to ordering more invasive UE evaluation. This enabled efficient testing and management of nearly 400 firefighters who might otherwise not have recognized their risk for BE, nor undergone any form of screening.

## Conclusion

This study capturing real-world data on the use of EsoGuard for early detection of BE/EAC in firefighters demonstrates its ability to test many individuals rapidly and efficiently in a well-tolerated fashion, and reliable use of the test to triage individuals prior to pursuing more invasive and time-consuming diagnostic approaches.

## Data Availability

All data produced in the present study are available upon reasonable request to the authors

